# Optimizing Spatial Distribution of Wastewater-Based Disease Surveillance to Advance Health Equity

**DOI:** 10.1101/2024.05.02.24306777

**Authors:** Maria L. Daza–Torres, J. Cricelio Montesinos-López, César Herrera, Yury E. García, Colleen C. Naughton, Heather N. Bischel, Miriam Nuño

## Abstract

In 2022, the US Centers for Disease Control and Prevention commissioned the National Academies of Sciences, Engineering, and Medicine to assess the role of community-level wastewater-based disease surveillance (WDS) beyond COVID-19. WDS is recognized as a promising mechanism for promptly identifying infectious diseases, including COVID-19 and other novel pathogens. An important conclusion drawn from this initiative is that it is crucial to maintain equity and expand access to maximize the advantages of WDS for marginalized communities. To address this need, we propose an optimization framework that focuses on the strategic allocation of wastewater monitoring resources at the wastewater treatment plant level. The framework’s purpose is to obtain a balanced spatial distribution, inclusive population coverage, and efficient representation of vulnerable communities in allocating resources for WDS. This study offers an opportunity to improve wastewater surveillance by tailoring location selection strategies to address specific priorities, improving decision-making in public health responses.

## 1 Introduction

Wastewater-based disease surveillance (WDS) is a rapidly evolving field that involves the measurement of health-related biomarkers excreted by individuals into sewer systems. WDS data offers real-time estimation of public health indicators and early warning of community disease outbreaks. Initially used to monitor poliovirus and detect pharmaceuticals and illicit drugs [1, 2], its utility greatly expanded during the Coronavirus Disease 19 (COVID-19) pandemic [3]. Widespread implementation of WDS during the pandemic proved to be a cost-effective method to monitor the dynamics of Sars Coronavirus 2 (SARS-CoV-2) community spread [4].

To enhance the nation’s ability to monitor SARS-CoV-2 effectively, the US Centers for Disease Control and Prevention (CDC) established the National Wastewater Surveillance System (NWSS) in September 2020. The NWSS expanded to encompass 1,154 testing sites nationwide as of March 2024. These sites collect samples from wastewater systems serving approximately 128 million people across the United States [5]. A critical factor behind the success of WDS is the widespread coverage of municipal wastewater collection systems, which link approximately eighty percent of households across the United States [6]. The NWSS is consolidating disparate local initiatives into a resilient and enduring national surveillance network.

WDS data can aid public health mitigation strategies by providing critical insights into the prevalence of COVID-19 within a community [5]. However, additional research is still needed to effectively implement WDS programs [7, 8, 9, 10]. A WDS program must possess sufficient spatial and temporal resolution to offer timely and dependable detection and alerts regarding disease outbreaks. Sampling sites are predominantly selected from wastewater treatment plants (WWTPs) or sewer maintenance holes, contingent on factors such as the size and demographics of the target population, the level of risk in the area, and the laboratory’s resources and capabilities.

Temporal and spatial resolution should undergo regular reassessment to ensure the system can detect significant changes with an adequate lead time to inform public health action effectively. The current distribution of sampling sites within the NWSS primarily comprises localities, tribes, and states that volunteered to participate during the pandemic emergency. This distribution might not accurately reflect the diverse demographic and geographic characteristics essential for establishing a comprehensive national network [11]. Furthermore, it may lack equity, optimal actionability, and long-term sustainability [12].

Several recent studies proposed optimal algorithms for selecting sewer monitoring sites to detect infectious diseases or hotspots at a catchment level [13, 14, 15, 16]. These algorithms aim to maximize the sensitivity of pathogen detection in the sewer network by considering factors like shedding, loss, decay, transport, and the population of the infectious agent. They utilize graph theory and optimization algorithms to identify hotspots and zero-patient locations within a sewer network. These algorithms are often complex and computationally intensive, which may limit their applicability to large-scale or national-level WDS programs. In this study, we introduce an optimization framework designed to strategically allocate WDS resources at the WWTP level to guide a WDS program. The framework takes into account a range of factors that may influence the effectiveness and efficiency of WDS. These include spatial distribution, population coverage, the presence of vulnerable communities, population density, and variability in wastewater signals. The optimization objective is to maximize a weighted combination of these factors while minimizing the total number of WWTPs actively monitoring wastewater. We address this challenge as an integer optimization problem and utilize the simulated annealing (SA) algorithm for an efficient solution. We assess various scenarios by adjusting the weightings for each factor to optimize the configurations of WWTPs in the WDS system. The primary objective is to minimize the total number of WWTPs actively involved in WDS.

This approach optimizes resource allocation in WDS by offering tailored location selection strategies that effectively address specific priorities and scenarios. By streamlining WDS efforts and reducing the number of WWTPs collecting samples, we can expand WDS to new communities, especially those at high risk, thereby broadening the scope of our efforts. This not only enhances the efficiency and cost-effectiveness of WDS but also ensures equitable support for informed decision-making across all communities.

## 2 Data description

We built the analytical framework using publicly accessible data in California, USA. We obtained data from the California Department of Public Health (CDPH) for 84 WWTPs that have been monitored for SARS-CoV-2 [17]. We include the following variables in our analysis: 1)*Geographic coordinates*, which include longitude and latitude of each WWTP based on its zip code location, as reported in [17]; this geo-location data is utilized to potentially reduce the selection of WWTPs close to each other, promoting a more geographically dispersed distribution, 2) *Population served*, representing the estimated number of individuals served by each sampling site; the incorporation of this term in our analysis prioritizes WWTPs that serve larger populations [17], 3) *Population density* measures individuals per square mile for each city where a WWTP is located [18]; we use population density data to prioritize areas with high population density, which may be more susceptible to disease transmission, 4) *Social Vulnerability Index*, which uses 16 US census variables in a relative measure of a population’s risk during a public health due to external stressors on health such as socioeconomic status and demographic factors [19]. We incorporate the social vulnerability index (SVI) at a county level to ensure the representation of vulnerable populations in the optimization problem. This factor is essential, particularly in regions with insufficient infrastructure and limited resources, which tend to be disproportionately affected during pandemics.

### SARS-CoV-2 RNA concentrations

We analyzed wastewater concentration data from 19 of the 84 WWTPs that monitored wastewater in California between March 21, 2022, and May 21, 2023. We selected this period because it corresponds with the availability of published protocols for sampling, analysis, and data processing, as outlined in the Supplementary Material (SM). Wastewater data were obtained from CPDH and WastewaterSCAN [17, 20]. The concentration of SARS-CoV-2 RNA measured in each wastewater sample was normalized by the concentration of pepper mild mottle virus (PMMoV) RNA measured in the same sample. PMMoV is a fecal indicator and laboratory process control that is commonly measured alongside SARS-CoV-2 to help mitigate noise arising from analytical processes, population size, and wastewater flow variations. Normalization results in the dimensionless metric N/PMMoV, which we refer to as the normalized wastewater concentration.

Based on this normalized wastewater concentration data, we construct a dissimilarity matrix that measures the difference between the wastewater signals of different WWTPs based on their patterns, such as trends, peaks, and variations. Figure 1A illustrates the spatial distribution of 84 WWTPs monitoring for SARS-CoV-2, along with the population they serve and SVI at the county level. The subset of 19 WWTPs utilized in the optimization problem involving the factor of signal dissimilarity is illustrated in Figure 4A.

**Figure 1:**
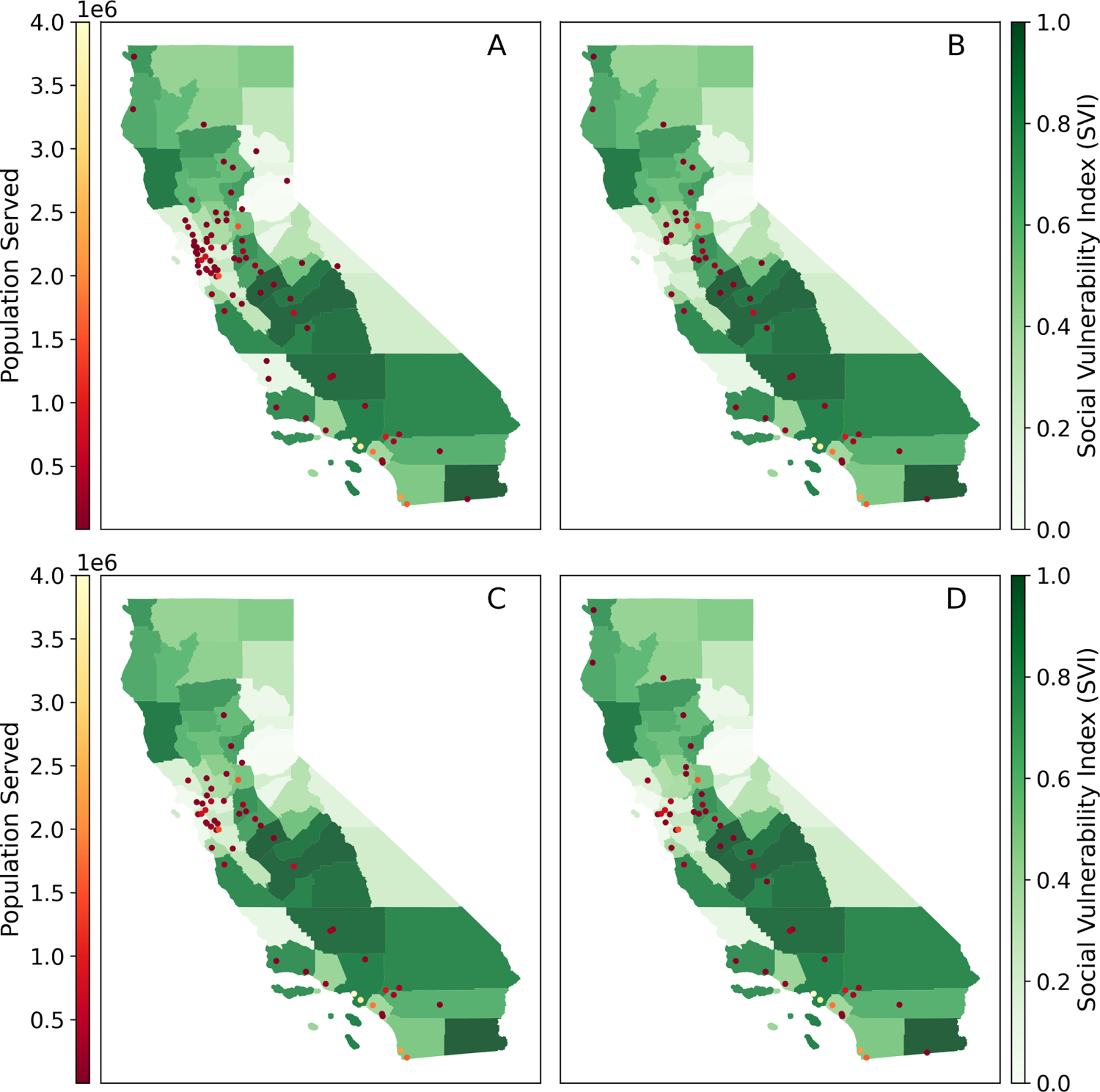
**A.** Geographic distribution of WWTPs across California monitoring SARS-CoV-2, with emphasis on the population served and SVI at the county level. Optimal solutions assuming a 40% reduction of active plants and different scenarios: **B.** Prioritizing SVI, **C.** Prioritizing Population Served, and **D.** Balanced approach.

## 3 Methods

This study aims to inform WDS planning in California by considering spatial distributions of WWTPs, population coverage, representation of vulnerable populations, and dissimilarity between wastewater signals. We normalize the factors to a common scale to facilitate comparisons, and we assign weights to the factors to demonstrate a range of strategies for balancing their relative importance. We then maximize the weighted combinations of factors while satisfying pre-defined constraints to minimize the number of WWTPs. This approach acknowledges practical considerations and constraints associated with wastewater monitoring capabilities.

### 3.1 Optimization problem formulation

Let *I* = *{*1*, …, n}* represent the set of WWTPs to be optimized. The objective function, denoted as *f* (x), seeks to maximize a weighted combination of various factors:

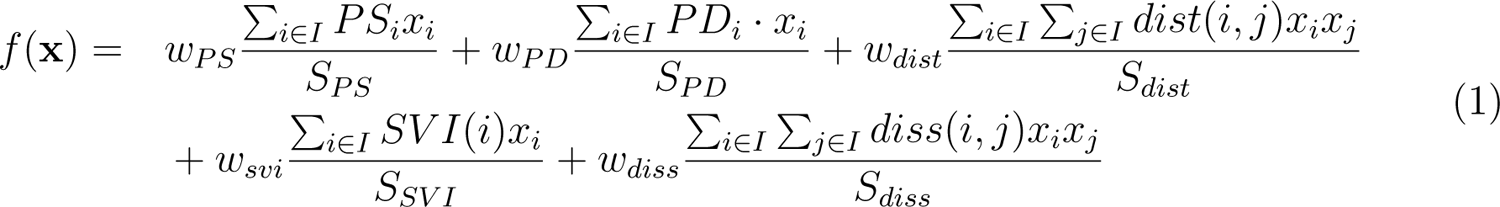

where x = (*x*_1_*, …, x_n_*) and *x_i_* is a binary variable indicating if plant *i* is selected (*x_i_*= 1) or not (*x_i_* = 0). The terms *PS_i_*, *PD_i_*, *dist*(*i, j*), *SV I*(*i*), and *diss*(*i, j*) correspond to the population served by plant *i*, city’s population density associated with plant *i*, the distance between plant *i* and plant *j*, SVI of the county associated with plant *i*, and dissimilarity between WWTP signals, respectively.

The variables *S_PS_* = Σ*_i∈I_ PS_i_*, *S_PD_* = Σ*_i∈I_ PD_i_*, *S_dist_* = Σ*_i∈I_* Σ*_j∈I_ dist*(*i, j*), *S_SV I_* = *Σ_i∈I_ SV I*(*i*), and *S_diss_* = *Σ_i∈I_ Σ_j∈I_ diss*(*i, j*) represent the total values of the corresponding terms over all plants. These sums facilitate normalization for comparison and the introduction of weights. The weights *w_PS_*, *w_PD_*, *w_dist_*, *w_SV I_*, and *w_diss_* assigned to each term in the objective function determine the importance given to the respective factors during the optimization process. To ensure balance, the sum of weights is constrained to be equal to 1: w*_PS_* + w*_PD_* + w*_dist_* + w*_SV I_* + w*_diss_* = 1.

The optimization problem is subject to the constraint that the total number of selected plants should be equal to a predefined number *k* (i.e., Σ*_i∈I_ x_i_* = *k*). This constraint plays a critical role in maintaining the efficiency and effectiveness of the WDS system, especially when dealing with resource limitations. The goal of the optimization problem is to select a subset of *k* WWTPs that effectively balance multiple factors mentioned above.

### 3.2 Computing a dissimilarity matrix using wavelets

Epidemiological studies often involve analyzing time series data to extract valuable insights and patterns across different scales. However, epidemiological processes are typically not stationary, implying that their statistical properties vary over time. Hence, a global timescale decomposition, where the entire time series is treated uniformly, may not be suitable for studying epidemiological processes [21]. Wavelet analysis is a suitable method for investigating non-stationary time series data, as it performs a local timescale decomposition of the signal [22]. Additionally, it allows us to measure the associations between two or more time series at any frequency band and time window, allowing us to detect synchrony, phase, and coherence patterns [21, 23]. It has been applied to various human infectious diseases, such as measles, influenza, leishmaniasis, and dengue [23, 24, 25, 26].

#### 3.2.1 Dissimilarity matrix

We apply the discrete wavelet transform (DWT) to the SARS-CoV-2 concentration time series data using the Daubechies 4 wavelet family. Decomposing the data into four levels (*L* = 4), this process yields coefficients at different scales and positions. At each level (*l*), we obtain approximation coefficients (*cA_l_*) and detail coefficients (*cD_l_*), both of length *N/*(2*^l^*), where *N* is the length of the wastewater time series. *cA_l_* contains information about the overall signal trend, while *cD_l_* contains high-frequency information or details, *l* = 1*, …, L*. These wavelet coefficients capture the characteristics of the signal at different scales and positions in both the time and frequency domains. We use these coefficients, excluding detail coefficients at the first two levels, to compute a dissimilarity matrix that measures how different the time series are based on their wavelet coefficients; see details in SM.

We used the correlation distance as the dissimilarity measure. The correlation distance between *u* and *v* is defined as 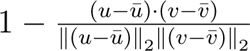, where ū is the mean of the elements of u and *x · y* is the dot product of *x* and *y*. A low correlation distance (high positive correlation) indicates a low dissimilarity, and vice versa. Other distance measures, such as the Euclidean distance, may not be suitable for time series with different scales or magnitudes since it only considers the absolute difference between two vectors. Time series with similar patterns but distinct scales could yield high dissimilarity with Euclidean distance, while correlation distance would give a low dissimilarity. Therefore, correlation distance may be better for comparing time series with different scales or magnitudes.

We use the PyWavelets library in Python to perform the DWT, and subsequently compute the correlation distance matrix using the scikit-learn library. This matrix is denoted as *diss*(*·, ·*) in our optimization function. Refer to Figure 3 for a visual representation of the computed correlation distance matrix.

The primary concept behind using wavelet coefficients to compare signals is to emphasize the patterns and relative changes in the data across different scales. Wavelet coefficients are relative measures that describe the variations in the data at different scales, regardless of the absolute magnitude of the data. These coefficients are normalized in the sense that they quantify the relative importance of different frequency components in the data. When comparing wavelet coefficients from two-time series, we are essentially comparing how their frequency content and patterns vary across scales, which makes it possible to identify similarities or differences in their underlying structures, even if the original magnitudes of the series are different.

#### 3.2.2 Simulated annealing

Optimal location problems are solved through exact methods or heuristic approaches, depending on the problem’s size and complexity [27]. Exact methods find the global optimum but are only feasible for small problems. Heuristic approaches aim to find good-quality solutions within a limited amount of computation time or space [28]. We use the SA algorithm, a heuristic approach that has proven effective in solving complex combinatorial optimization problems [29].

SA is inspired by the annealing process in metallurgy, where a metal is heated quickly to a high “temperature” (a parameter) and then gradually cooled to a more ordered state. By gradually cooling down, SA facilitates effective exploration of the solution space and finds an optimal solution that meets our objectives. The “temperature” parameter in SA controls the balance between exploration (searching for new solutions) and exploitation (improving existing solutions). This algorithm reduces the risk of falling into local minima common to iterative improvement methods, as unlike the latter, it accepts solutions that worsen the objective function [28].

Let *I* = *{*1*, …, n}* denote the set of WWTPs to be optimized. Algorithm S1 in SM delineates the steps for maximizing the objective function *f* (x), given in Equation(1), using SA, while ensuring that the total number of selected plants equals *k < n*. To initialize the candidate solution x_0_, we randomly select *k* distinct numbers from *I*, forming a set of indices denoted as *I_k_*. The entries of x_0_ are set to 1 if their corresponding indices are in *I_k_*, and 0 otherwise. For generating a new candidate solution x_new_, we replace a randomly chosen number from *I_k_*with a new random number from *I−I_k_*(i.e., the set of elements in *I* but not in *I_k_*). Following the guidelines outlined in [30], we adapt the “temperature” based on whether we have accepted 10 *× n* solutions or generated 100 *× n* candidate solutions. In the reported results, we fix *T*_max_ = 10^5^ (maximum temperature), *T*_min_ = 10*^−^*^7^ (minimum temperature), and *α* = 0.1 (cooling schedule parameter). Then, we compute the energy in both the current and the new candidate solutions: *E* = *f* (x_0_) and *E*_new_ = *f* (x_new_), respectively. Then, we calculate the change in energy, Δ*E* = *E − E*_new_. If Δ*E <* 0, indicating an improvement, we accept the new solution x_new_. Otherwise, we generate a random number *r* from a uniform distribution *U* (0, 1). If *r < e^−^*^Δ*E/T*^, where *T* is the current temperature, we accept x_new_; otherwise, we reject it.

## 4 Results

We developed a flexible framework that can be used to evaluate resource planning scenarios, tailored to address specific challenges and priorities within a WDS system. We consider two overarching scenarios: 1) a disease-agnostic approach that considers only geographic and population attributes in resource allocation, and 2) a disease-informed approach that additionally considers dissimilarities in the dynamics of wastewater-derived COVID-19 data. The first scenario facilitates the prioritization of vulnerable populations using the SVI, while the second further facilitates the prioritization of unique wastewater signal contributions to the WDS dataset. In each scenario, we reduced the number of WWTPs from baseline by 40%. We explore three configurations within each scenario that align with potential system priorities, achieved by assigning varying weights to the parameters of the objective function. Sustainable, efficient, and equitable allocation of WDS resources is essential for the continued effectiveness of surveillance efforts in the long term. Our framework offers a strategy and tool for public health officials and policymakers involved in wastewater-based disease surveillance and resource allocation.

### Scenario 1: Disease-agnostic optimization approach

In Scenario 1, we integrated data on the location and population served by 84 WWTPs in California as the baseline condition, with city-level population density and the county-level SVI associated with each facility. We assigned a weight of zero to the normalized wastewater concentration dissimilarity term in the optimization function, representing scenarios for which disease dynamics are not considered in site selection. Reducing the number of plants by 40% resulted in a total of 50 remaining WWTPs in each solution. We explored three configurations of weight combinations to gain an understanding of how weights influence the solution:

**– Scenario 1A: Prioritizing SVI (***w_SV I_* = 1*, w_PS_* = *w_PD_* = *w_dist_* = *w_diss_* = 0). This scenario prioritizes plants with high SVI, aiming to improve representation in areas with increased social vulnerabilities. The optimization solution (see Figure 1B) prioritizes the selection of plants situated in areas with higher SVI scores, which tend to correspond to communities that experienced disproportionate impacts from COVID-19.

**– Scenario 1B: Prioritizing population served (***w_PS_* = 1*, w_PD_* = *w_dist_* = *w_SV I_* = *w_diss_* = 0). Prioritizing population served directs attention to ensuring that surveillance efforts are concentrated in areas where a larger number of people reside (*w_PS_* = 1) (see Figure 1C).

**– Scenario 1C: A balanced approach (***w_PS_* = *w_PD_* = *w_dist_* = *w_SV I_* = *w_diss_* = 0.25*, w_diss_* = 0). This scenario seeks an optimal solution that balances multiple factors equally (see Figure 1D).

Findings in Figure 1 underscore key insights essential for determining the allocation of disease surveillance resources. In the scenario prioritizing SVI, numerous plants in California’s Central Coast region are omitted (Figure 1B). However, plants in the Central Valley and South Interior regions are retained, which aligns with the high SVI scores observed in those areas. This highlights a potential trade-off: while prioritizing SVI helps address vulnerable communities, it might lead to gaps in geographic coverage. In the scenario prioritizing population served (Figure 1C), two plants in the North region, one in the South Coast, and several in the Central region of California are eliminated. Communities represented by some of these plants correspond to regions in California that experienced a significant burden during the COVID-19 pandemic. The removal of these plants could have important implications for public health and environmental equity, particularly in communities that already have limited access to healthcare and socioeconomic disparities. The allocation of WWTPs guided by a balanced set of priorities, as described in Figure 1D, seems to provide reasonable results despite the significant downsizing of plants.

Figure 2 describes the distribution of SVI, population served, and density among all 84 WWTPs included in this scenario. As expected, scenarios prioritizing specific factors such as SVI and population served display higher values and less variability in these variables compared to all plants. Interestingly, a balanced approach consistently results in the selection of WWTPs with similar distributions for the population served and density while also representing communities with higher SVI scores. Notably, the two plants that ceased data collection consistently appear as optimal solutions in multiple scenarios. One plant, located in the Central Valley, emerged as an optimal solution across all scenarios, while the other, located in a Southern County of CA, was optimal in the SVI and balanced priority scenarios.

**Figure 2:**
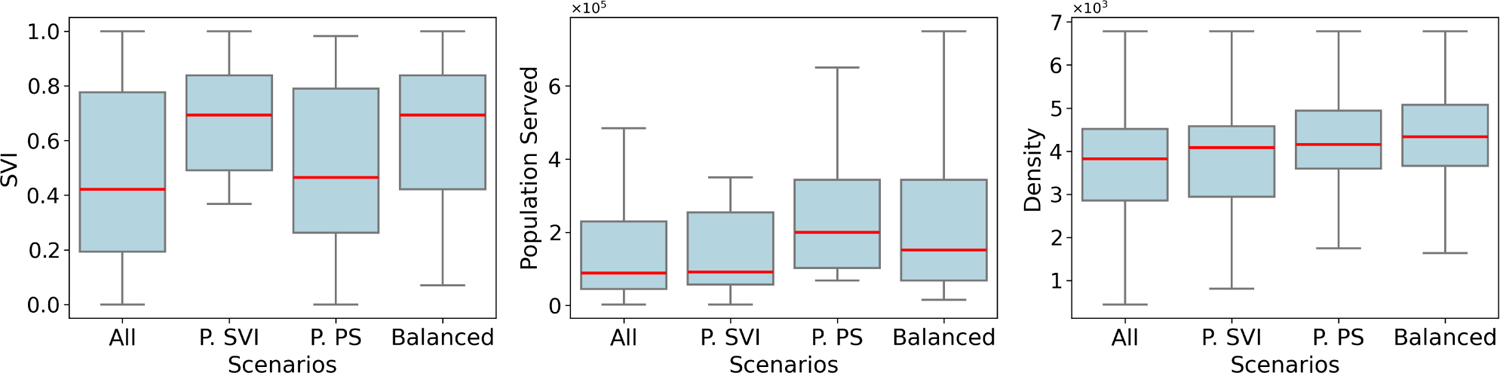
Distribution of the SVI, population served, and population density for all WWTPs that have WDS for SARS-CoV-2 in California (All), and for the optimal WWTPs found in each scenario: prioritizing SVI (P.SVI), prioritizing population served (P.PS), and a balanced approach (Balanced). Outliers were excluded from the boxplot for improved visualization; see Figure S.5 in SM for a plot including outliers.

### Scenario 2: Disease-informed optimization approach

Scenario 2 represents a planning scenario in which the dynamics of the community disease trends are considered alongside geographic and population factors in the selection of sampling sites. We include a dissimilarity term that takes into account the differences between normalized wastewater concentrations, giving preference to those with distinct signal patterns. We consider 19 WWTPs as a baseline in the scenario with a wastewater signal dissimilarity term in the optimization problem in Equation (1).The dissimilarity matrix, which represents the correlation distance between discrete wavelet transforms of normalized SARS-CoV-2 wastewater concentrations, is illustrated in Figure 3. By incorporating dissimilarity term in the objective function, our goal is to present the results of the optimization problem and demonstrate the impact of this addition.

**Figure 3:**
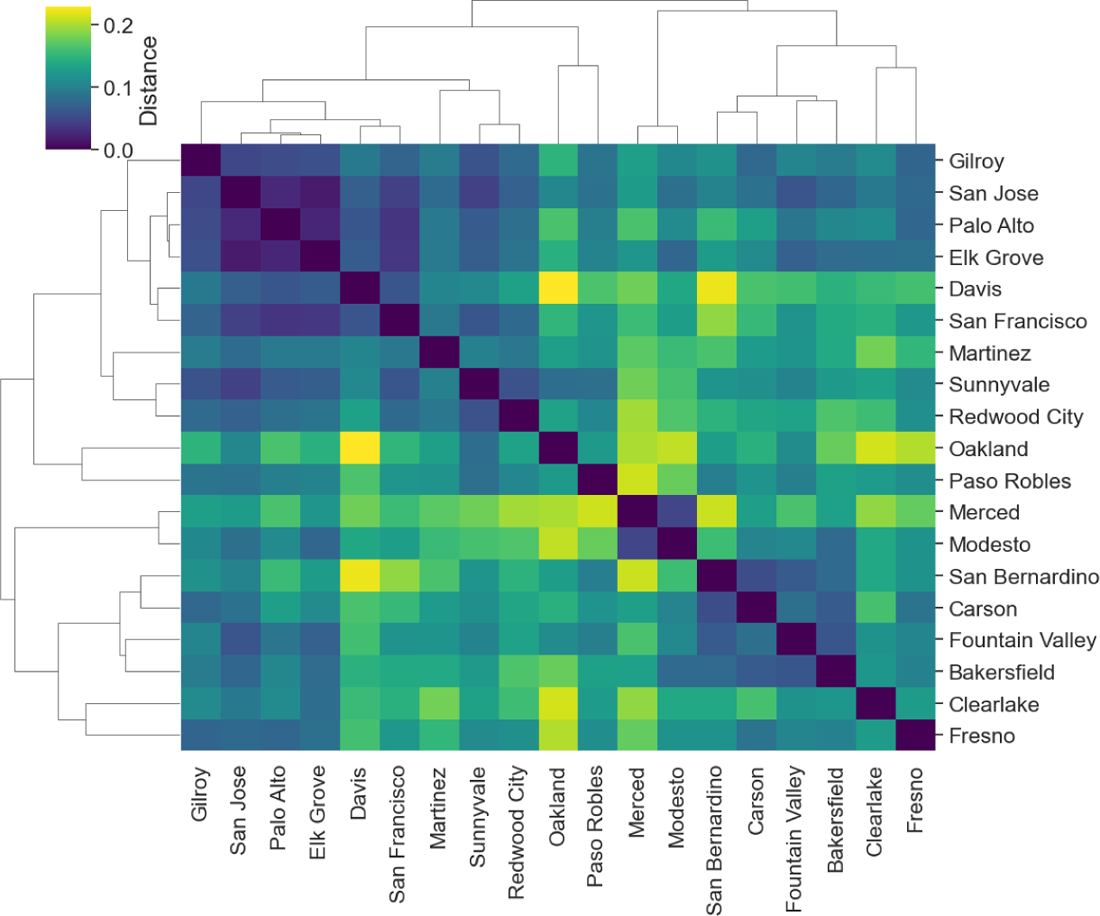
Dissimilarity matrix depicting wavelet coefficients for 19 WWTPs. Alongside the matrix, dendrograms illustrate hierarchical clustering, showcasing how WWTPs group together based on similarities in wavelet coefficient.

**Figure 4:**
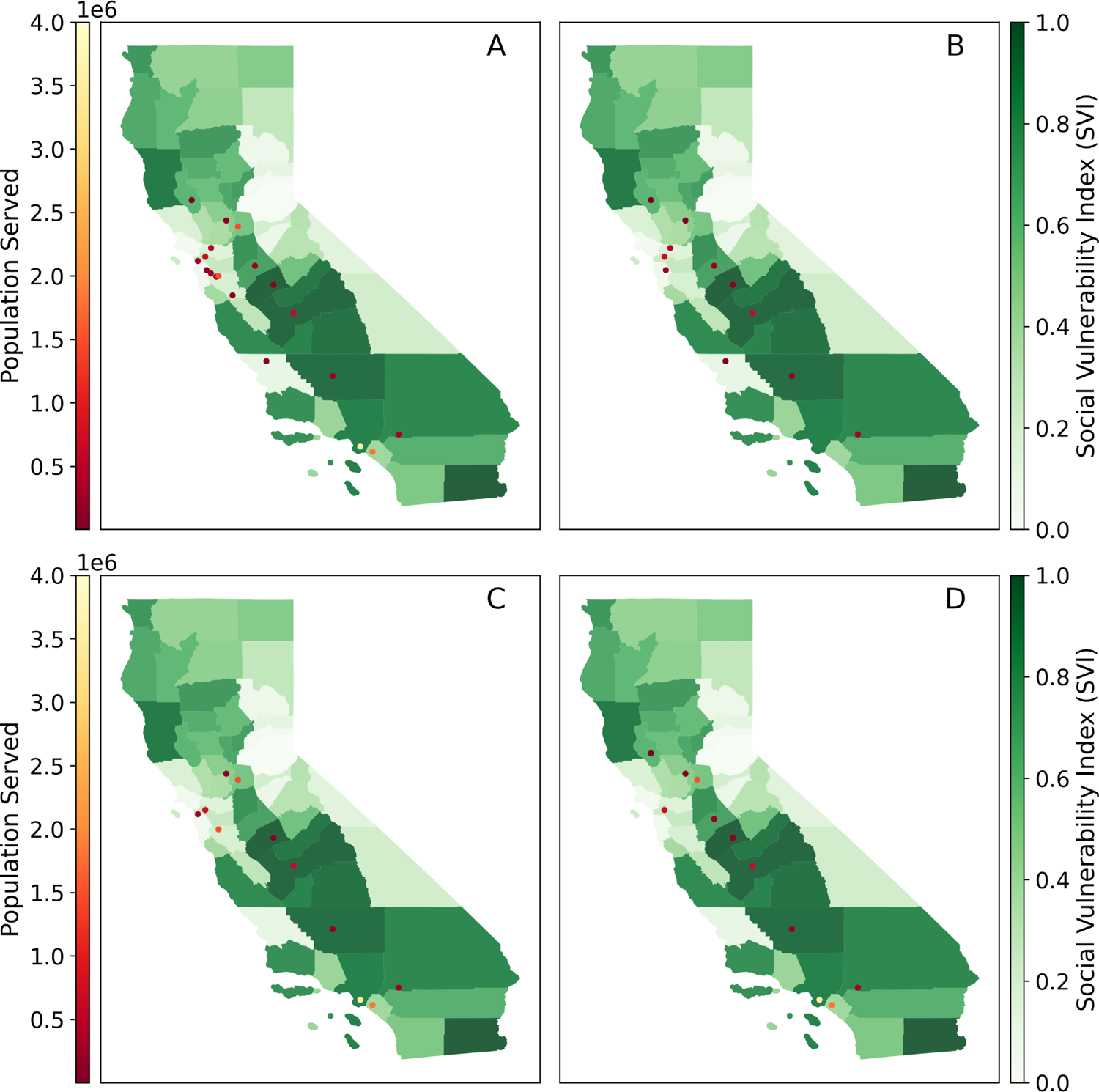
Optimal solutions obtained assuming signal dissimilarity and 19 initial WWTPs. Each priority scenario assumes a goal of a 40% reduction in the number of plants included in the optimization problem. **A.** Initial Plants. **B.** Prioritizing disease signal dissimilarity. **C.** Equal factors. **D.** A balanced approach for SVI and disease signal dissimilarity.

**– Scenario 2A: Prioritizing disease signal dissimilarity (***w_diss_* = 1*, w_PS_* = *w_PD_* = *w_dist_* = *w_SV I_* = 0). This scenario emphasizes dissimilarity between WDS signals, aiming to optimize the representation of unique signal patterns across WWTPs (see Figure 4A).

**– Scenario 2B: Equal factors (***w_PS_* = *w_PD_* = *w_dist_* = *w_SV I_* = *w_diss_* = 0.2). This scenario seeks an optimal solution that weights multiple factors equally (see Figure 4B).

**– Scenario 2C: A balanced approach for SVI and disease signal dissimilarity (***w_SV I_* = *w_diss_*= 0.35*, w_PS_*= *w_PD_* = *w_dist_* = 0.1). In this scenario, we emphasize both the SVI and dissimilarity metrics, while also taking into account population density, spatial distances, and population served (see Figure 4C).

Prioritizing dissimilarities in our optimization process can improve our capacity for early detection, response, and tailored interventions. Figure 4 illustrates the WWTPs selected in the optimization problem prioritizing WW signal dissimilarity (Figure 4B), initially with 19 plants (Figure 4A). Subsequently, it showcases the plants that remained after adopting an equal factors approach (Figure 4C) and a balanced approach for SVI and disease signal dissimilarity (Figure 4D).

Figure S.4 in SM displays the dissimilarity matrix for each optimal solution and scenario. In this particular scenario, prioritizing the dissimilarity term means that we are placing emphasis on highlighting the distinctions between the selected WWTPs with regard to their wastewater signal characteristics.

## 5 Discussion

In this paper, we present an optimization framework to evaluate scenarios for the selection of WWTPs for wastewater-based disease surveillance. Our analysis in California revealed that prioritizing social vulnerability in defining the optimization problem enables more equitable integration of communities with a history of significant COVID-19 burden when planning the allocation of resources for WDS. These findings can inform public health surveillance efforts, particularly as WDS is adapted to help manage infectious diseases beyond COVID-19, including for influenza and respiratory syncytial virus. By focusing on areas with higher vulnerability, which are often characterized by limited resources and greater health disparities, resources for surveillance efforts can be allocated more effectively. This strategy helps ensure that communities with elevated risks of infection and severe symptoms receive greater attention and support in disease monitoring and response.

Our findings make a significant contribution to the current literature by bringing to light the complexities of optimizing WDS resource allocation. We demonstrate how prioritizing key factors in the optimization process could influence planning decisions for resource distributions. Our results also highlight how different communities offer unique contributions to a WDS program. For instance, we observed that two WWTPs within the sampling frame ceased WDS data collection during the study period, yet these two facilities were consistently retained in most of our optimization scenarios. Our framework offers a mechanism to evaluate the significance of WDS program changes in the context of seeking equitable representation and cost efficiencies. We recommend ongoing evaluation of site distributions to maintain fair and inclusive coverage. Greater investments in community engagement and partnership-building programs can support marginalized communities and guarantee they have a say in decision-making processes related to WDS.

Actively addressing equity in WDS planning and resource allocation is crucial to strengthening pandemic preparedness. One solution is to adopt a more comprehensive strategy that gives priority to vulnerable communities in the distribution of monitoring resources. This could entail re-evaluating the standards used to choose monitoring sites to ensure sufficient coverage of regions with elevated social vulnerability. Our approach is optimized from the 84 WWTPs that have had WDS due to data availability. The scenarios examined in this study serve as valuable entry points, offering initial insights into potential WWTP configurations, but there are over 200 WWTPs in California that could be further evaluated in the future. The framework can also be extended to other states and countries to guide resource allocation for WDS. It can be customized and adjusted to ensure that surveillance efforts are fair, efficient, and in line with the changing requirements of diverse communities, ultimately enhancing our collective capacity to detect and mitigate public health risks on a global level. While this framework provides valuable insights and serves as a basis for decision-making, we recognize several limitations and needs for further improvement. First, achieving more practical and context-specific outcomes will require accounting for decision-maker preferences and constraints in future refinements. For instance, budget limitations, workforce availability, and infrastructure capacity can be incorporated to inform proposed solutions. Second, reliability and accuracy of the optimization model are reliant on the quality and availability of data pertaining to population distribution, social vulnerability, historical disease burden, and other related variables. Enhancing data transparency and accessibility can empower communities to participate actively in WDS efforts and advocate for their requirements.

Finally, integrating spatial and temporal dynamics into upcoming optimization models could provide significant improvements to the modeling framework. The distribution of population and disease burden may vary spatially and temporally, requiring dynamic modeling approaches to account for these fluctuations accurately. Furthermore, ethical considerations and other equity concerns should also be factored into the optimization process to guarantee a fair and unbiased allocation of disease surveillance resources [31, 32]. To this end, it is essential to establish clear guidelines and criteria that prioritize the needs of under-served and marginalized communities. This may involve conducting thorough assessments of socio-economic factors, healthcare access, and vulnerable indices to identify areas with the greatest need for surveillance resources. Moreover, stakeholder engagement and community involvement are crucial components of ensuring equity in resource allocation. By actively seeking input from affected communities, local leaders, and advocacy groups, decision-makers can gain valuable insights into the specific challenges and priorities of different population groups.

## Data Availability

All data produced in the present work are contained in the manuscript.

## Acknowledgments

This research was conducted as part of Healthy Central Valley Together (HCVT), a collaborative research project to enable public health action from wastewater-based disease surveillance. Research support for HCVT was provided by a philanthropic gift to the University of California, Davis. We extend our gratitude to the California Department of Public Health (CDPH) and WastewaterSCAN for their collaboration and for generously sharing wastewater data. We would like to express our gratitude to Alexandria B. Boehm, Marlene K. Wolfe, and Jesus De Loera for their valuable comments. We extend our sincere gratitude to the dedicated teams at the wastewater treatment plants, as well as the invaluable support from public health entities and all other collaborators involved in the collection and curation of the data used in this study.

## Supplementary Material

## S1 Comparative analysis of wastewater data using discrete wavelet transform

### S1.1 Wavelet decomposition

The wavelet transform decomposes non-stationary time series into a time-frequency domain, allowing interpretation of temporal variability. It analyzes signals at multiple resolutions and scales, using basis functions called wavelets. Wavelets are mathematical functions used to decompose data into different frequencies and scales. The continuous wavelet transform (CWT) is given by the following equation:

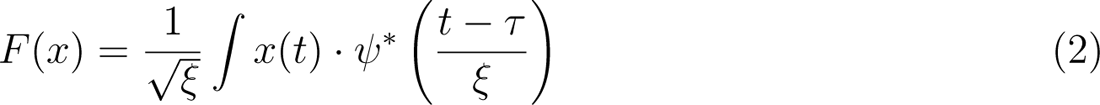

where *F* (*x*) is the wavelet transform for the signal *x*(*t*) as a function of time, *ξ* is the scale parameter, *τ* is the time parameter, and *ψ* is the mother wavelet or the basis function with *∗* denoting the complex conjugate.

The CWT is computationally expensive, but the discrete wavelet transform (DWT) offers efficiency. DWT uses sub-band coding, which is a technique of dividing the signal into frequency bands or sub-bands, and then sampling each sub-band at a lower rate. The DWT achieves this by passing the signal through a series of high-pass and low-pass filters, which produce detail and approximation coefficients, respectively. The approximation coefficients estimate the rough features of the original data, while the detail coefficients capture frequent movements, such as sudden changes or spikes, that might be hidden in the original data due to trends or seasonal fluctuations. The DWT facilitates multi-resolution representations of signals, aiding pattern detection algorithms by revealing structures at various time scales.

### S1.2 Comparative analysis

In this paper, we used data from two sources, the California Department of Public Health (CPDH) and WastewaterSCAN (WWSCAN) [17, 20], each of which implemented different sampling and laboratory analytical protocols to measure the concentration of SARS-CoV-2 RNA (N gene) and PMMoV (Tables S.2-S.3). The CDPH laboratory analyzed raw wastewater from composite influent samples, with concentrations of each gene target reported as genome copies per volume of wastewater. The WWSCAN laboratory analyzed dewatered settled solids from composite influent samples or, more often, dewatered solids from primary clarifier sludge, with concentrations of each gene target reported as genomic copies per mass of dry sludge. Trends in normalized wastewater concentrations (N/PMMoV, a dimensionless metric) obtained by the two methods have been extensively validated against trends COVID-19 clinical data. However, the magnitudes of the normalized wastewater concentrations differ between the procedures. Distinct laboratory procedures can also introduce differing noise, biases, or variations into the data, which may manifest as rapid fluctuations or high-frequency components.

A primary challenge thus lies in directly comparing wastewater signals acquired by different methods due to the inherent biases introduced by different laboratory procedures. To help mitigate these biases, we employed a wavelet transform, which extracts patterns and features from the data to compare and analyze the wastewater signals more robustly. Specifically, we decompose the data into multiple low-frequency and high-frequency components, allowing us to focus independently on high-frequency details. We hypothesize that variations in high-frequency components may arise from differences in laboratory methodologies rather than fluctuations in disease dynamics. We conduct an exploratory analysis to investigate the success of a wavelet transformation in distinguishing patterns related to disease dynamics rather than features specific to the laboratory methods.

We analyzed normalized wastewater concentration data collected from the city of Davis between May 7, 2022, and September 28, 2022, by two different laboratories applying highly similar methods (referred to as Lab 1 and Lab 2). For a detailed description of this data, please refer to [33]. We used the normalized wastewater concentration of SARS-CoV-2 RNA detected in each wastewater sample for the two laboratories. We employed the discrete wavelet transformation utilizing the Daubechies 4 wavelet family to decompose the data into multiple levels. During this process, coefficients at different scales and positions are generated.

At each level (*l*), we obtain two sets of coefficients: approximation coefficients (*cA_l_*), representing low-frequency components) and detail coefficients (*cD_l_*), representing high-frequency components). Both *cA_l_* and *cD_l_* have a length of *N/*2*^l^*. *cA_l_* captures information about the overall trend of the signal, while *cD_l_* contains high-frequency details, *l* = 1, *… L*.

We utilized the PyWavelets library for the wavelet decomposition, specifically the pywt.wavedec function. We employed the pywt.dwt_max_level to determine the appropriate decomposition level function. The pywt.wavedec function returns the coefficients (*cA*4, *cD*1, *cD*2, *cD*3, *cD*4). Figure S.1 shows the wavelet coefficients for the two-time series. We observed higher differences between these two series in the detail coefficients *cD*_2_, *cD*_3_.

**Figure S.1:**
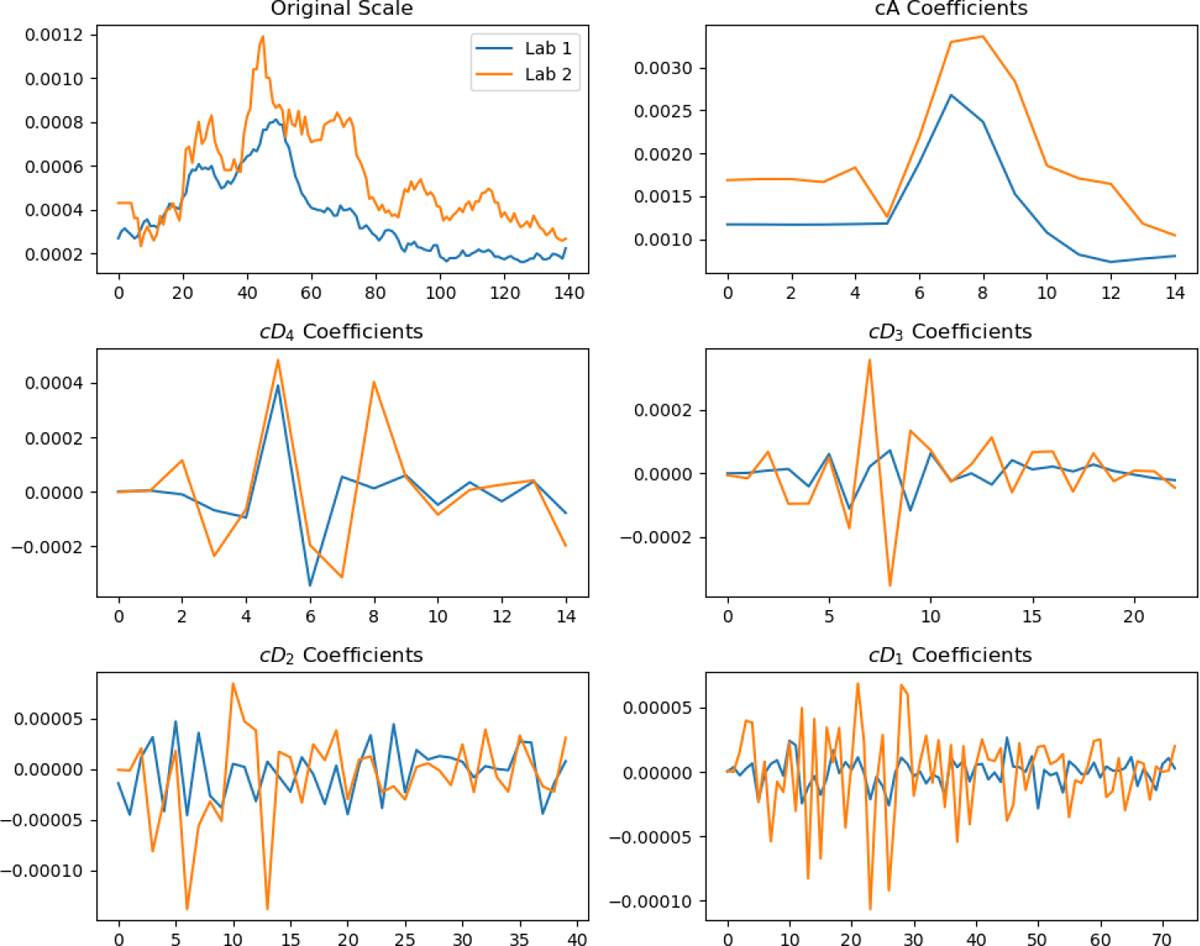
Wavelet Coefficients Comparison between Lab 1 and Lab 2 time series. We calculated the distance correlation between the data from the two labs using different data representations: original scale, wavelet coefficients (including all levels), and wavelet coefficients excluding detail coefficients at different levels. The results, as shown in Table S.1, indicate that the distance correlation is lower (indicating higher similarity) when we use their wavelet coefficients instead of the series in the original scale. We also observed that excluding detail coefficients at the first two levels further decreased the distance correlation, as these coefficients primarily capture changes in high frequencies within the data and may be associated with laboratory-specific methods.

**Table S.1:**
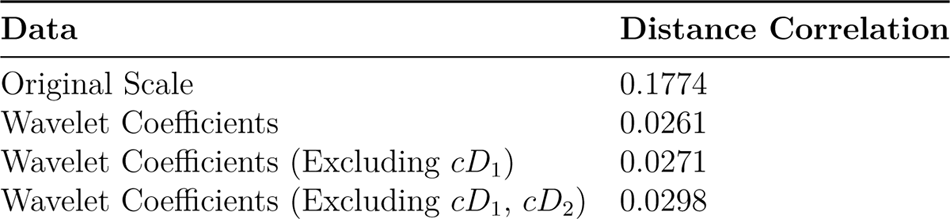
Distance correlation between Lab 1 and Lab 2.

| Data | Distance Correlation |
| --- | --- |
| Original Scale | 0.1774 |
| Wavelet Coefficients | 0.0261 |
| Wavelet Coefficients (Excluding $cD_1$) | 0.0271 |
| Wavelet Coefficients (Excluding $cD_1, cD_2$) | 0.0298 |

Furthermore, we reconstructed the time series from the wavelet coefficients, excluding coefficients at level 1, levels 1 and 2, and levels 1, 2, and 3. When we exclude detail coefficients at level 1 and at levels 1 and 2, the reconstructed signal closely aligns with the original series, with only small variations removed, Figure S.2. However, excluding detail coefficients at the third level results in a desynchronization of the reconstructed signal compared to the original scales, leading to a loss of information about the trend. This highlights the importance of preserving essential features while eliminating high-frequency components during signal reconstruction.

**Figure S.2:**
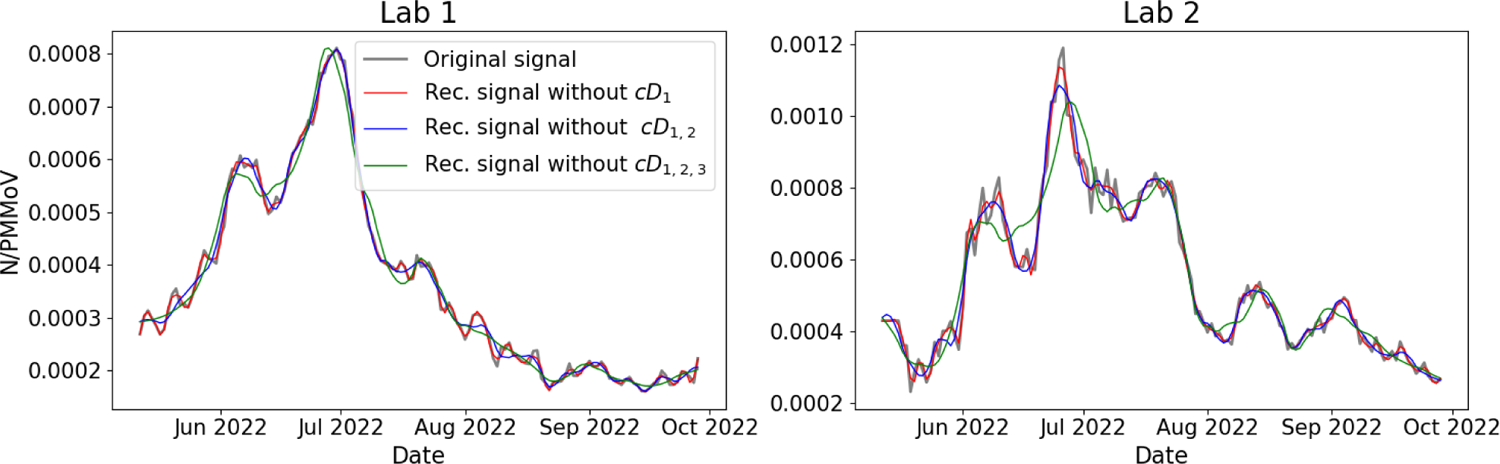
Reconstructed time series from wavelet coefficients excluding different levels.

Based on our findings, we chose to use wavelet coefficients, excluding detail coefficients at levels 1 and 2, to construct the dissimilarity matrix with correlation distance. This approach helped us identify similarities between signals from different laboratories while minimizing the biases introduced by varying measurement methods. We will explore this hypothesis further in the future analysis to assess the variability in different frequencies and scales and its relation with different labs, procedures, temperature, and other variables.

## S2 Algorithm performance

To validate the robustness of the algorithm, we executed it 100 times, consistently converging to the same solution in each run. This demonstrates the stability and reliability of the algorithm across multiple iterations. Figure S.3 illustrates the energy trajectories for these runs, providing a visual representation of the algorithm’s convergence consistency.

**Figure S.3:**
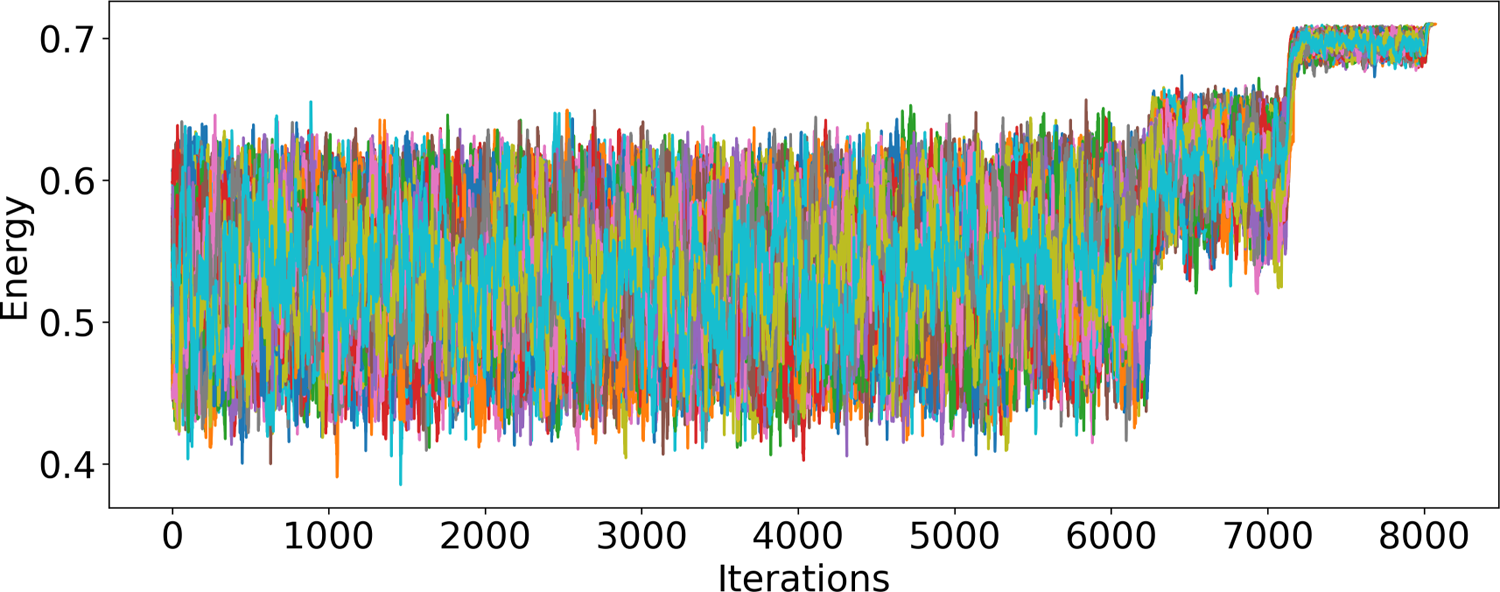
Evaluation of the optimization function is performed in each iteration of simulated annealing. The traces correspond to 100 trajectories that were run to demonstrate their performance.

## S3 Suplementary figures

**Figure S.4:**
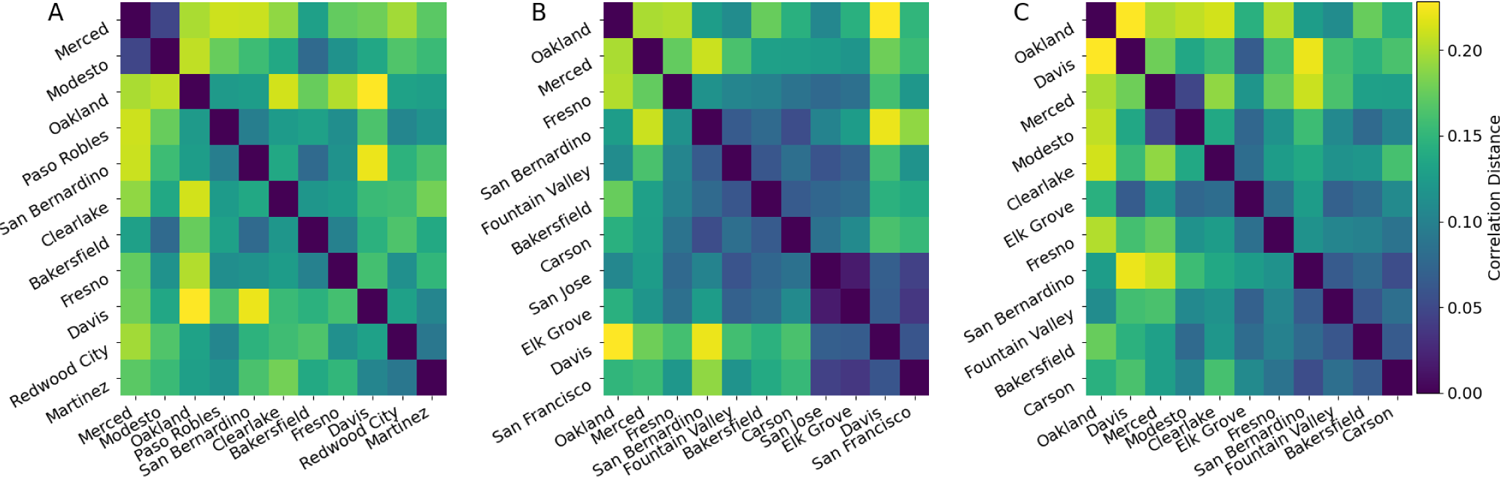
Dissimilarity matrix for the optimal solution in each scenario. **A.** Prioritizing disease signal dissimilarity. **B.** Equal factors. **C.** A balanced approach for SVI and disease signal dissimilarity.

**Figure S.5:**
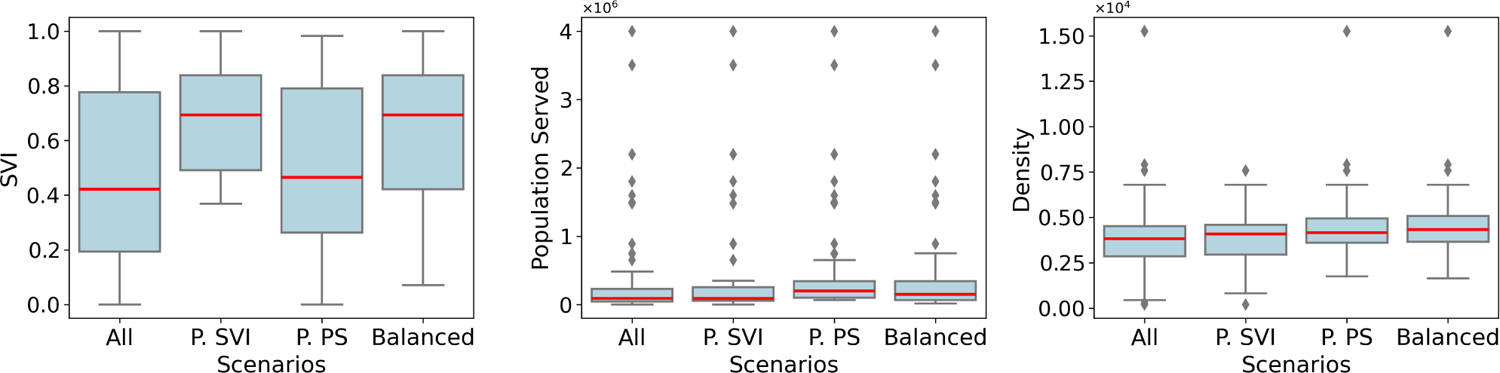
Distribution of the SVI, population served, and population density for all WWTPs that have WDS for SARS-CoV-2 in California (All), and for the optimal WWTPs found in each scenario: prioritizing SVI (P.SVI), prioritizing population served (P.PS), and a balanced approach (Balanced).

## S4 Simulated Annealing algorithm

**Algorithm 1.**
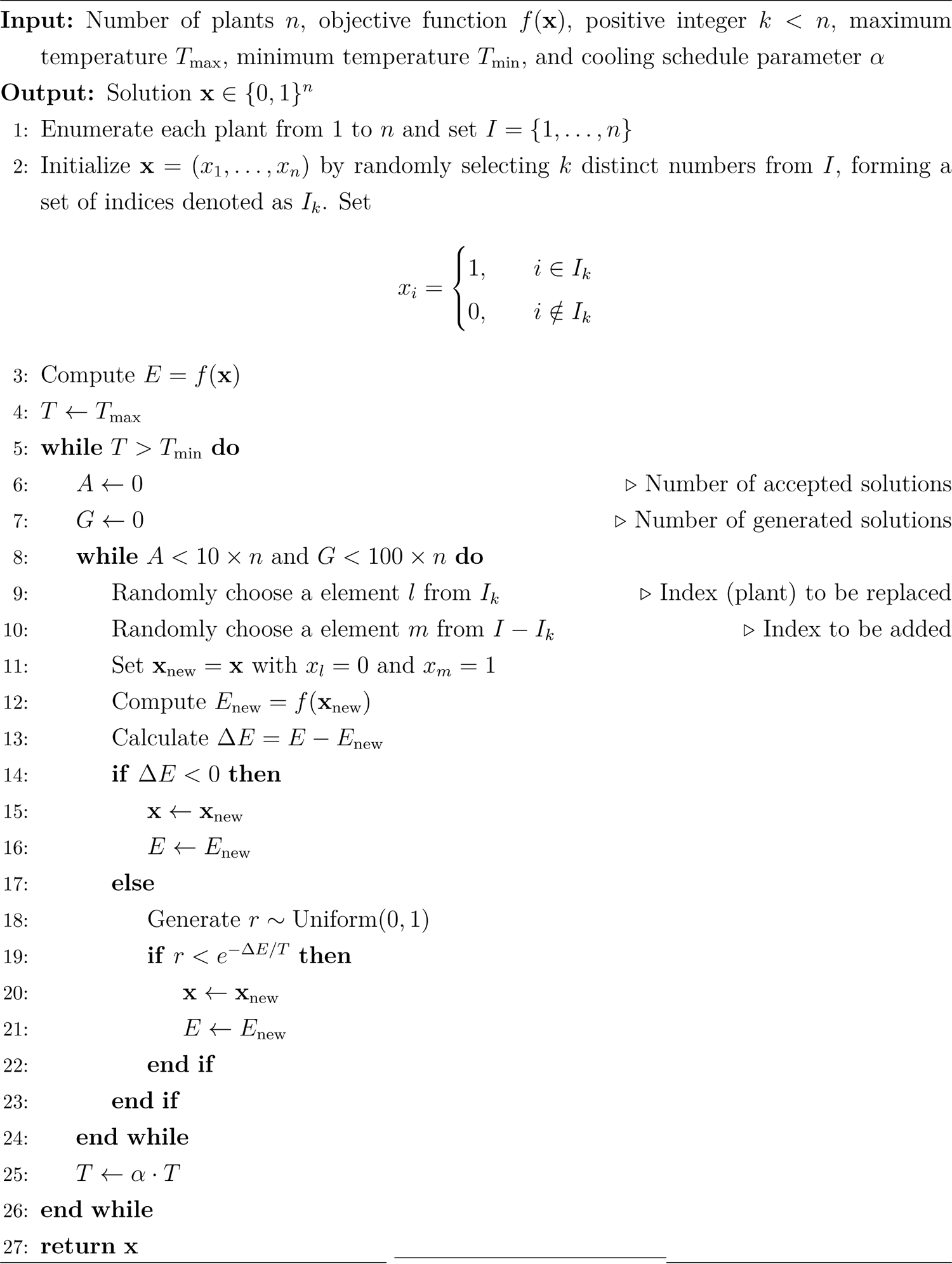
Simulated Annealing Algorithm

## S5 Wastewater methods description

Wastewater data from March 21, 2022, to May 21, 2023, for WWTPs located in Oakland (Alameda), Martinez (Contra Costa), Merced (Merced), Elk Grove (Sacramento), Paso Robles (San Luis Obispo), Palo Alto (San Mateo), San Jose (Santa Clara), Modesto (Stanislaus), and Davis (Yolo) have been previously published in [34]. For the remaining WWTPs, we present an overview of the methods employed as reported by CDPH. Specifically, we refer to the lab identifiers listed in Table S.3 for the WWTPs described in Table S.2.

**Table S.2:**
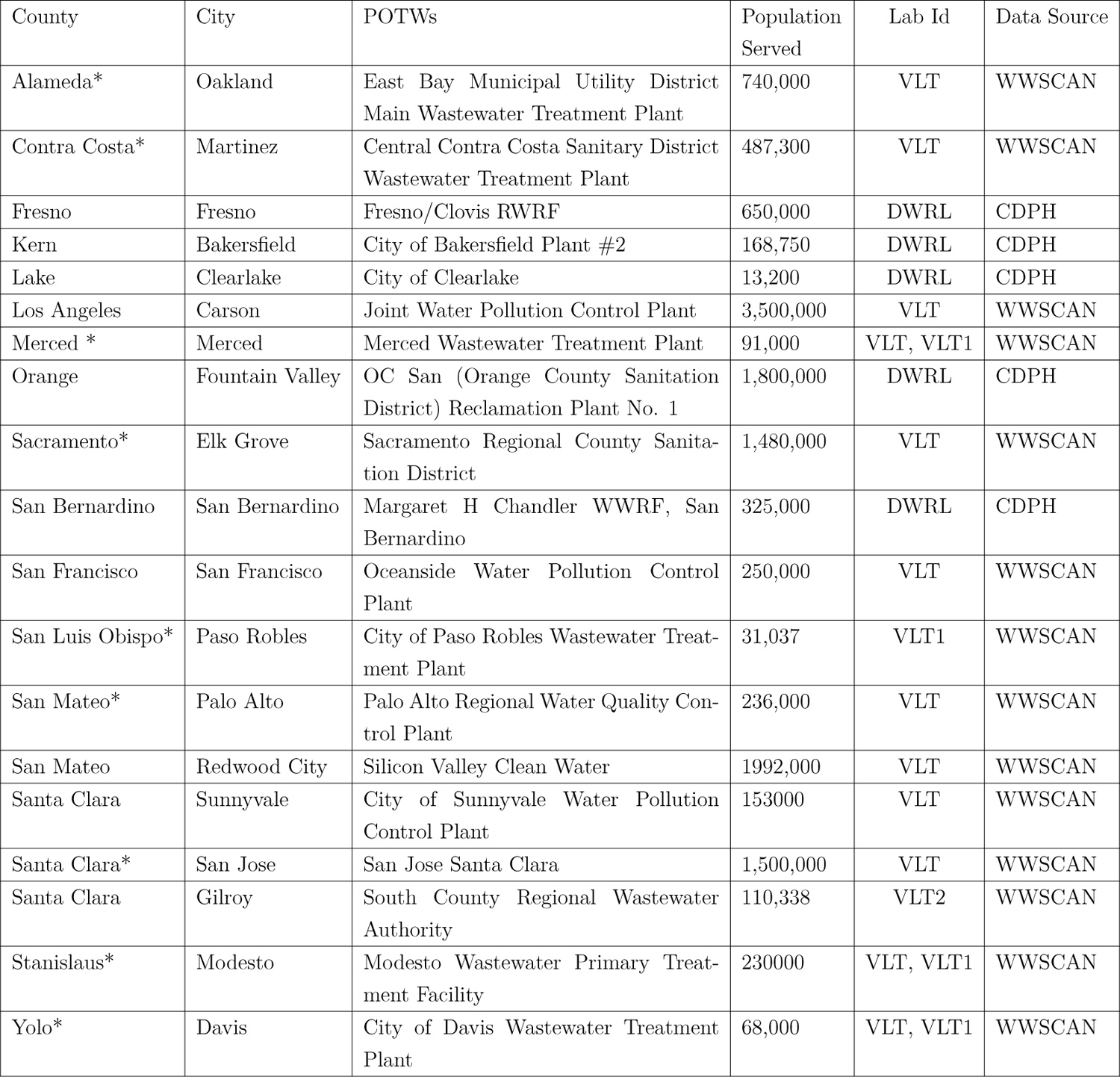
Description of publicly owned treatment works (POTWs) included in this study. Plants located in cities marked with an asterisk (*) indicate those for which wastewater data during the study period have been published here [34]. Lab Id (DWLR; VLT, VLT1, VLT2) corresponds to methods summarized in Table S.3.

**Table S.3:**
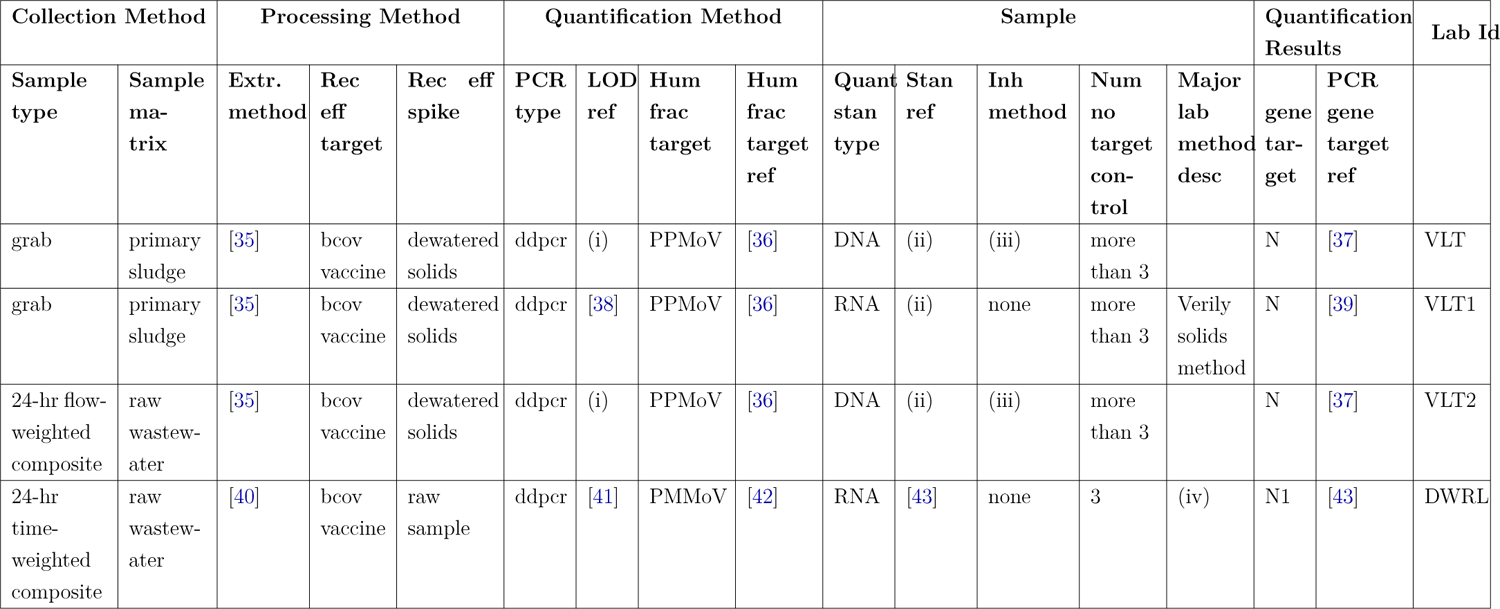
Summary of different methods employed in the collection, processing, quantification, and analysis of samples for the detection of viral pathogens in wastewater as reported by CDPH [17]. Abbreviations: Extr. method - Extraction method; Rec eff - Recovery efficiency; PCR - Polymerase Chain Reaction; LOD ref - Limit of detection reference; Hum frac - Human fecal fraction; Quant stan type - Quantification standard type; Stan ref - Standard reference; Inh method - Inhibition method; Lab - Laboratory; Major lab method desc - Major laboratory method description. References are provided where applicable.(i) 3 droplets as a minimum for a positive detection. (ii) Synthetic DNA oligo purchased from IDT. (iii) Solutions titrated with varying concentrations of solids to identify a concentration at which inhibition of the SARS-CoV-2 assays was minimized. (iv) This lab uses a lab method distinct from other labs in this reporting jurisdiction.

